# Improving outcomes for children with malaria, diarrhoea and pneumonia in Mozambique through the inSCALE technology innovation: A cluster randomised controlled trial

**DOI:** 10.1101/2022.07.25.22278035

**Authors:** Seyi Soremekun, Karin Källander, Raghu Lingam, Ana-Cristina Castel Branco, Neha Batura, Daniel Strachan, Abel Muiambo, Nelson Salomao, Juliao Condoane, Fenias Benhane, Frida Kasteng, Anna Vassall, Zelee Hill, Guus ten Asbroek, Sylvia Meek, James Tibenderana, Betty Kirkwood

**Affiliations:** Department of Infection Biology, London School of Hygiene & Tropical Medicine, Keppel Street, London, WC1E 7HT, United Kingdom; Malaria Consortium, The Green House, 244-254 Cambridge Heath Road, London, E2 9DA, United Kingdom; Department of Global Public Health, Karolinska Institutet, 17177, Stockholm, Sweden; UNICEF Programme Division, Health Section, New York, NY, United States; Population Child Health Research Group, School of Women’s and Children’s Health, University of New South Wales. Australia; Malaria Consortium, Rua Joseph Ki-Zerbo 191, PO Box 3655, Coop, Maputo, Mozambique; Institute for Global Health, University College London, 30 Guilford Street, London WC1N 1EH, United Kingdom; Department of Global Health and Development, London School of Hygiene & Tropical Medicine, Keppel Street, London, WC1E 7HT, United Kingdom; Institute for Global Health, University College London, 30 Guilford Street, London WC1N 1EH. United Kingdom; The Nossal Institute for Global Health, Melbourne School of Population and Global Health, The University of Melbourne Victoria, 3010, Australia; Department of Global Health, Amsterdam University Medical Centres, Meibergdreef 9 1105 AZ Amsterdam, The Netherlands; Amsterdam Institute for Global Health and Development, Paasheuvelweg 25, 1105 BP Amsterdam, The Netherlands; Department of Population Health, London School of Hygiene & Tropical Medicine, Keppel Street, London, WC1E 7HT, United Kingdom

## Abstract

**Background:** The majority of post-neonatal deaths in children under 5 are due to malaria, diarrhoea and pneumonia (MDP). The WHO recommends integrated community case management (iCCM) of these conditions using community-based health workers. However iCCM programmes have suffered from poor implementation and mixed outcomes. We designed and evaluated a technology-based intervention ‘inSCALE’ (<u>In</u>novations At <u>Sc</u>ale For <u>L</u>asting <u>E</u>ffects) to support iCCM programmes and increase appropriate treatment and other outcomes for children with MDP.

**Methods:** This superiority cluster randomised controlled trial allocated all 12 districts in Inhambane Province in Mozambique to receive iCCM only (control) or iCCM plus the inSCALE technology intervention. The key components of the intervention consisted of a digital application on smartphones and tablets providing clinical decision support algorithms, stock tracking, automatic personalised messaging, free calls, and solar chargers for iCCM-trained community health workers and primary care facility supervising staff in intervention districts. Population surveys were conducted at baseline and after 18 months in all districts to assess the impact of the intervention on the coverage of appropriate treatment for malaria, diarrhoea and pneumonia in children 2-59months of age, on prevalence of cases of these conditions, and on a range of secondary household and health worker level outcomes. All statistical models accounted for the clustered study design and variables used to constrain the randomisation. A meta-analysis of the estimated pooled impact of the technology intervention was conducted including results from a sister trial (inSCALE-Uganda).

**Findings:** The study included 2740 eligible children in control arm districts and 2863 children in intervention districts. The prevalence of cases of MDP decreased from 53.5% (1467) to 43.7% (1251) in the control and intervention arms respectively (risk ratio 0.82, 95% CI 0.78-0.87, p<0.001). The rate of care seeking to the iCCM-trained community health worker increased in the intervention arm (14.4% vs 15.9% in control and intervention arms respectively) but fell short of the significance threshold (adjusted RR 1.63, 95% CI 0.93-2.85, p=0.085). Coverage of the appropriate treatment of cases of MDP increased by 26% in the intervention arm (RR 1.26 95% CI 1.12-1.42, p<0.001) after accounting for the randomisation and design effects. Across two country trials, the estimated pooled effect of the inSCALE intervention on coverage of appropriate treatment for MDP was RR 1.15 (95% CI 1.08-1.24, p <0.001).

**Interpretation:** The inSCALE intervention led to a reduction in prevalence of MDP and an improvement in appropriate treatment when delivered at scale in Mozambique. The programme will be rolled out by the ministry of health to the entire national CHW and primary care network in 2022. This study highlights the potential value of a technology intervention aimed at strengthening iCCM systems to address the largest causes of childhood morbidity and mortality in sub-Saharan Africa.

**Author Summary:** The inSCALE cluster-randomised trial in Mozambique was part of a $10million project funded by the Bill and Melinda Gates Foundation to design and test innovative primary care interventions to improve health outcomes for children with malaria, diarrhoea and pneumonia (MDP), which together are the largest killers of children aged <5yrs. The study aimed to strengthen the primary health care system with a focus on community health workers, representing the most accessible level of care for many underserved populations.

We designed a technology-based intervention delivered using cheap smartphones. This intervention was based on mHealth principles and included basic AI to guide correct diagnosis and treatment of MDP, provided personalised feedback to health workers, and alerts to supervising health facilities on stock outs and data tracking. The study was implemented within the entire province of Inhambane, and districts were randomly assigned to the intervention or to continue with standard care (control). Compared to control districts, we observed significant reductions in the prevalences of MDP in children under 5 years (reductions of 20% for malaria, 34% for pneumonia, and 45% for diarrhoea) and an increase in appropriate treatment of any cases of MDP by 26% (of all cases MDP) and 40% (of all children) in the intervention districts.

As a result of this trial, the government of Mozambique incorporated the inSCALE intervention into its policy for child health services, and is in the process of scaling up the programme to all 8000+ community health workers across the country (2022).

## Introduction

More than 5 million mostly preventable deaths occur in children under 5 years of age every year(1). The vast majority of post neonatal deaths in this group are due to pneumonia, diarrhoea, malaria or malnutrition(2). It is acknowledged that urgent work is required to address the causes and more widely, the systems within which such deaths occur. However progress has been uneven; under 5 mortality rates in sub Saharan Africa today remain similar to or higher than 1990 mortality rates in most high income countries with an average rate of 78 deaths per 1000 live births across the continent in 2018(2). Lack of access to timely and appropriate clinical management of sick children is a key factor driving poor health outcomes(3,4) prompting a World Health Organisation/UNICEF joint statement in 2012 advocating for integrated community case management (iCCM) of malaria, diarrhoea and pneumonia delivered by community-based lay health workers, who would be trained to diagnose and treat key childhood conditions in areas with poor access to facility-based services (3). In the decade following this statement, iCCM programmes have been introduced into more than 50 low- and middle-income countries (LMICs)(5–7). Whilst a recent multi-country modelling study in DRC, Malawi, Niger, and Nigeria (8) estimated that iCCM may have been responsible for averting nearly 5000 deaths per year in these countries combined due to malaria, diarrhoea and pneumonia, this estimate varied considerably between countries, and was in contrast to a Cochrane systematic review which found no clear impacts of iCCM on morbidity and mortality(9). Both studies reported poor or uneven implementation of iCCM programmes and a need for further studies that addressed structural deficiencies in health systems and processes that hamper success and scale-up.

Central to the challenge in implementation of iCCM has been the lack of sufficient support for community-based health worker (CHW) networks, low rates of CHW attrition, of reported motivation, and of quality of disease management practices(10). The WHO calls for “innovative and tailored approaches” to strengthening child health services (1), yet there are few programmes or even research studies which have explicitly addressed structural barriers in the context of scaling up iCCM effectively.

Digital technologies, and specifically mHealth (the use of mobile technologies in medical and public health practice (11) is an area of growing interest for the delivery of health services in sub-Saharan Africa (12,13) particularly given the rapid expansion of mobile phone networks and usage in this setting (14). In this paper we report on the evaluation of a systems-strengthening package of programmes collectively called “inSCALE” (<u>in</u>terventions at <u>s</u>cale for <u>c</u>ommunity <u>a</u>ccess and lasting <u>e</u>ffects). inSCALE was a multi-country study of innovative interventions implemented by the Malaria Consortium in partnership with the governments of Mozambique and Uganda and supported by international funders (the Bill and Melinda Gates Foundation, UNICEF, and the Canadian international Development Agency now Global Affairs Canada (GAC)). Reported in this paper is findings of the “inSCALE technology intervention”, a disease management support package that included electronic clinical decision algorithms, personalised reports, messaging, and feedback to CHWs and health facility staff. The Mozambique site tested the inSCALE technology intervention only; the Uganda site tested both the technology intervention and a community-based intervention described elsewhere [**Uganda ref**]. The technology interventions in both countries sought to specifically address primary health system challenges that could strengthen provision for key childhood illnesses through improving CHW performance, motivation and reducing attrition. The primary aim of the trial was to assess the impact of the inSCALE technology intervention on the coverage of the appropriate treatment of childhood malaria, diarrhoea and pneumonia. This paper reports on the impacts of the technology intervention in Mozambique to accompany the separate impact analysis for the Uganda inSCALE trial [**ref**] and conducts a meta-analysis of pooled impact of the technology intervention across its sites in Uganda and Mozambique.

## Results

In total 9231 households in the 12 districts were assessed for eligibility and of these, 495 were empty or no one was home, and 4483 did not contain children under 5 years of age. Overall, the main caretaker was available in 92% (3920) of houses with children under 5 years of age. At the endline survey, a total of 2718 children had had symptoms of MDP in the past month (the impact analysis sample), of which 1467 were in the control arm and 1251 in the intervention arm (Figure 1 flow chart). Household characteristics of sick children were fairly well balanced between arms with small variations in some parameters – Table 1.

**Table 1.** Characteristics of the inSCALE Mozambique study sample

| Characteristic | Control % (n) | Technology % (n) |
| --- | --- | --- |
| **Eligible children** | **N = 2740** | **N = 2863** |
| % children with MDP | 53.5% (1467) | 43.7% (1251) |
| % children with suspected malaria* | 48.5% (1300) | 39.8% (1118) |
| % children with confirmed malaria* | 24.6% (661) | 21.8% (612) |
| % children with diarrhoea** | 7.2% (198) | 4.2% (121) |
| % children with suspected pneumonia** | 16.2% (443) | 10.6% (304) |
| **Children with MDP** | **N = 1467** | **N = 1251** |
| % Respondent is mother of sick child | 81% (1181) | 83% (1036) |
| % children are boys | 50% (733) | 50% (629) |
| **Age of mother** |  |  |
| 12-19 | 10% (144) | 8% (104) |
| 20-29 | 38% (552) | 37% (467) |
| 30-39 | 28% (413) | 30% (373) |
| 40-49 | 12% (181) | 14% (175) |
| 50+ | 8% (113) | 6% (76) |
| Not known | 4% (64) | 4% (56) |
| **Highest education level completed (respondent)** |  |  |
| None | 31% (453) | 32% (400) |
| Incomplete primary | 48% (706) | 45% (557) |
| Primary or higher | 21% (308) | 24% (294) |
| **Mother tongue** |  |  |
| Bitonga | 17% (246) | 6% (81) |
| Chitsua | 54% (793) | 72% (900) |
| Chichopi | 17% (254) | 19% (242) |
| Other | 12% (174) | 2% (28) |
| **Marital status** |  |  |
| Married | 2% (34) | 3% (41) |
| Living with partner (not married) | 78% (1151) | 74% (929) |
| Single/divorced/widowed | 19% (282) | 22% (281) |
| **% Main occupation is farming/fishing** | **75.9% (1113)** | **64.0% (801)** |

\*Denominator is the 5490 children aged 4months – 5 years; \*\*Denominator is the 5603 children aged 2m – 5 year

**Figure 1.**
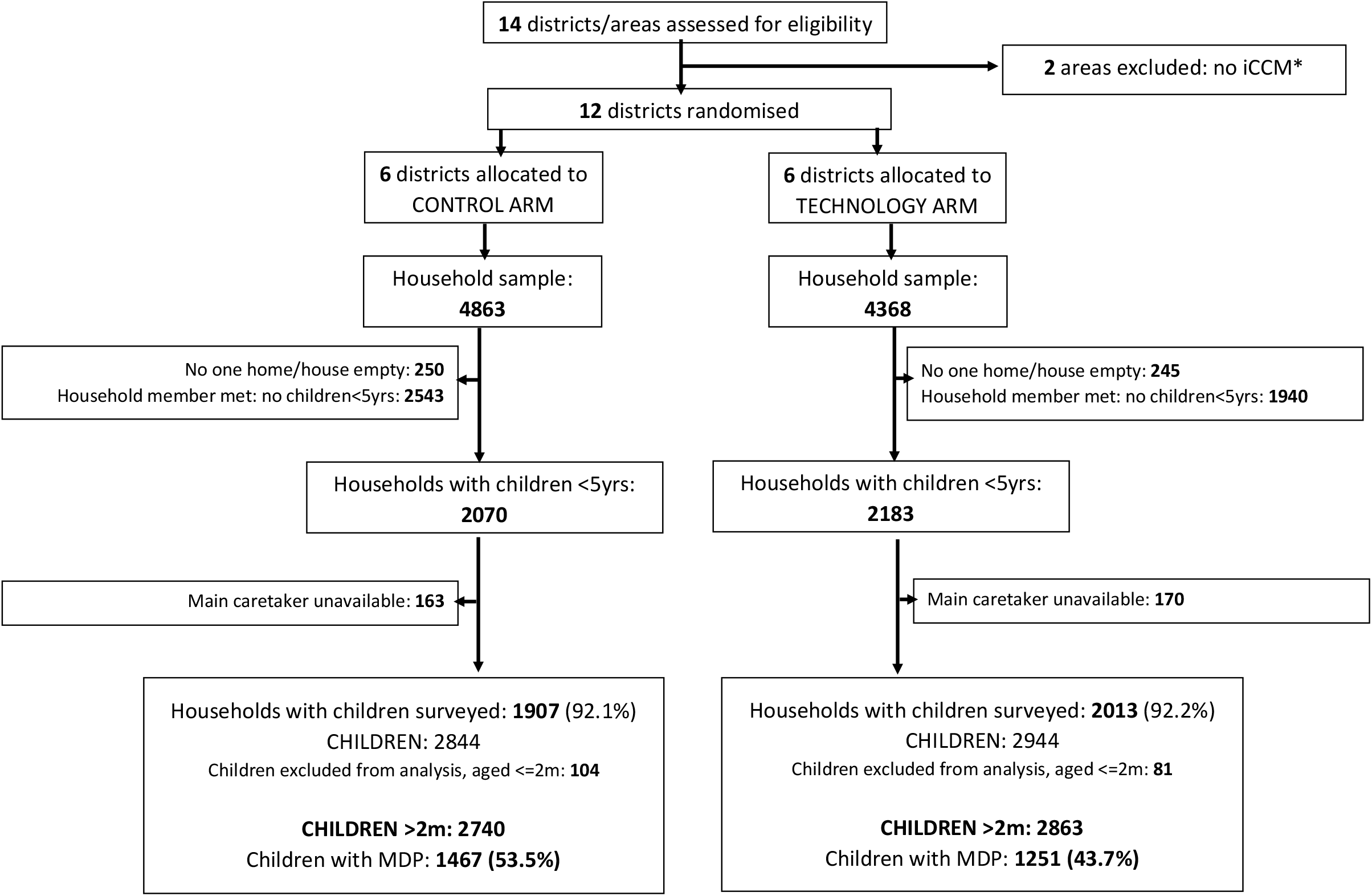
Flow diagram of participant progression through trial phases. Final analysis sample: children aged 2 months to 5 years with suspected malaria, diarrhoea or suspected pneumonia (MDP). * Maxixe and Inhambane, the economic and provincial capitals were excluded from the study (do not implement iCCM).

The restricted randomisation method resulted in good balance between intervention arms for balance variables as described in the protocol paper(15), which were care seeking rates, CHW motivation, and cost of seeking care (Table 2). Differences for all variables was either less than 5% (percentages), or less than 0.1 (means).

**Table 2.** Key indicators used to balance arms at baseline

| Variables included in the restricted randomisation* | Control arm<br>662 children with MDP<br>XX CHWs | Technology<br>658 children with MDP<br>XX CHWs | Difference |
| --- | --- | --- | --- |
| Mean (log<sub>10</sub>) cost of care seeking for children with MDP (SD) | 1.11 (0.09) | 1.14 (0.14) | 0.03 |
| Cluster mean % care seeking to an CHW for children with MDP (SD) | 22.5% (23.03) | 22.8% (16.92) | 0.26 |
| Cluster mean % care seeking to a public facility for children with MDP (SD) | 50.96% (21.63) | 48.79% (14.80) | 2.17 |
| Cluster mean CHW motivation score (SD) | 2.29 (0.46) | 2.38 (0.50) | 0.09 |

\*Outcomes are presented as mean percentages or mean costs across all clusters in each arm

### Intervention coverage, sickness and care seeking

By the end of the 18-month implementation period, 68% of CHWs reported having a working inSCALE phone and app, and 45% had sent a report via the app in the last 4 weeks (Table 3). Whilst overall prevalence of MDP was similar between arms at baseline, there were significant reductions in the prevalence of suspected malaria, diarrhoea and pneumonia (reductions ranging from 18% to 45%), and a 16% increase in the prevalence of diagnostic testing for malaria in the intervention arm at endline (Table 4). Care seeking to the CHW as the first port of call was 14.4% in the control arm and 15.9% in the intervention arm (199/1251). After accounting for the restricted randomisation and clustered design this represented a 63% increase in care-seeking to the CHW, a result that just fell short of significance (Table 5, p=0.085). There were no significant differences in care seeking rates to other providers or in the prevalence of children for whom no care was sought (Table 5).

**Table 3.**
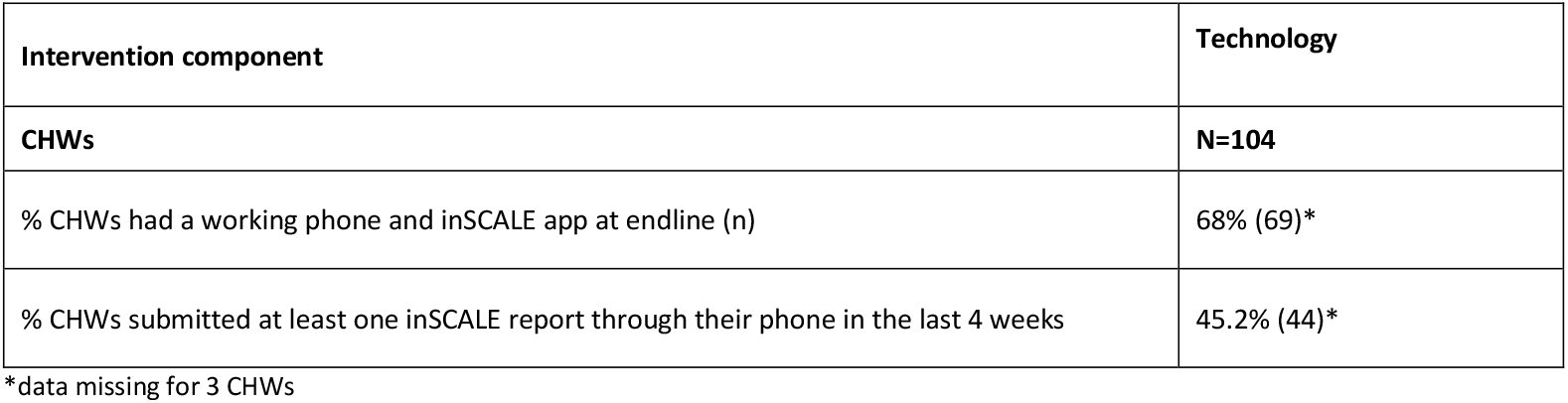
Exposure to intervention components recorded during the endline survey

**Table 4.** Prevalence of suspected malaria, diarrhoea and pneumonia (MDP) and blood tests for malaria in children under 5 years at baseline and endline

| Prevalence of | Baseline<br>(reported prevalence in last 2 weeks) |  | Endline<br>(reported prevalence in last 4 weeks) |  | Impact of inSCALE at endline<br>Risk ratio inSCALE/control<br>RR: 95% CI; p* |
| --- | --- | --- | --- | --- | --- |
|  | Control arm | Technology arm | Control arm | Technology arm |  |
| Any of MDP | 29.3% (662/2257) | 30.4% (658/2165) | 53.5% (1467/2740) | 43.7% (1251/2863) | **0.82** (0.78-0.87) p<0.001 |
| Suspected malaria* | 25.9% (562/2171) | 26.7% (556/2084) | 48.5% (1300/2682) | 39.8% (1118/2808) | **0.80** (0.72-0.89) p<0.001 |
| Blood test for malaria** | 31.7% (200/631) | 29.5% (181/613) | 59.7% (937/1570) | 62.3% (831/1335) | **1.16** (1.06-1.28) p=0.002 |
| Confirmed malaria*** | 65.5% (131/200) | 68.5% (124/181) | 70.5% (661/937) | 73.7% (612/831) | **1.04** (0.92-1.18) p=0.514 |
| Diarrhoea | 5.1% (116/2257) | 5.3% (114/2165) | 7.2% (198/2740) | 4.2% (121/2863) | **0.55** (0.42-0.71) p<0.001 |
| Pneumonia | 9.9% (223) | 11.8% (256/2165) | 16.2% (443/2740) | 10.6% (304/2863) | **0.66** (0.44-0.99) p=0.045 |

Prevalence (n/N) calculated as total eligible children 2 months-59 months of age (N) with condition in question (n), with the exception of fever and malaria where age range is 4 months-59 months in line with treatment guidelines. \*Fever, excludes fevers with a negative blood test for malaria. \*\*denominator includes all children reporting fever in the last two (baseline) or 4 (endline) weeks. \*\*\*percentage of all blood tested fevers that were positive. \*Adjusted for i) parameters used to balance arms at baseline and ii) the baseline prevalence of prevalence parameter

**Table 5.** Care seeking choices for children with MDP at endline

| Outcome % of children (n) | Control (N=1467) | Technology (N=1251) | Risk ratio tech/control<br>RR: 95% CI | p |
| --- | --- | --- | --- | --- |
| % seeking care at a CHW (first point of call) | 14.4% (211) | 15.9% (199) | 1.63 (0.93-2.85) | 0.085 |
| % seeking care at a CHW (at any point) | 14.7% (215) | 16.2% (203) | 1.61 (0.91-2.85) | 0.104 |
| % Not seeking care outside the home | 18.3% (269) | 20.6% (251) | 0.98 (0.74-1.30) | 0.898 |
| % seeking care at a public facility (at any point) | 67.4% (988) | 63.7% (797) | 1.11 (0.93-1.33) | 0.253 |
| % seeking care in the private sector (at any point - private facility, pharmacy, shop, herbalist etc) | 3.6% (53) | 2.4% (30) | 0.94 (0.51-1.71) | 0.839 |

### Appropriate Treatment

Baseline levels of appropriate treatment were similar for the combined MDP endpoint but varied by provider and for individual conditions between intervention arms (Table 6b). At endline there was an 26% increase in appropriate treatment of cases of MDP (the primary outcome (Table 6a) in the inSCALE arm compared to the control arm (95% confidence interval: 12%-42%, p<0.001). This rose to 30% when only episodes treated with recommended first line treatments were defined as appropriately treated (Table 6a 95% CI: 12%-52%, p=0.001). A breakdown of appropriate treatment by provider suggests this impact was driven by significant improvements in appropriate treatment coverage in public health facilities (Table 6a) which were the first port of call for caregivers for more than 60% of sick children in both arms (Table 5), and in the appropriate treatment of diarrhoea, where a 76% increase (95% CI 20%-256% p=0.003) in the use of oral rehydration salts was observed in the intervention arm (Table 7). Note it was not possible to estimate a confidence interval for use of ORS plus zinc due to small cell sizes. Additional improvements came in the use of the recommended first line antibiotic for pneumonia (50% increase in prescribing of amoxicillin in the intervention arm 95% CI: 3%-217%). A whole-child analysis (Supplementary File 1 Table S6) estimated an improvement of 40% in the proportion of children appropriately treated for all MDP condition(s) reported in the last 4 weeks in the intervention arm (95% CI: 9% - 79% p=0.009).

**Table 6a.** Appropriate treatment: Appropriate treatment of episodes suspected malaria, diarrhoea or suspected pneumonia (MDP)

| Appropriate treatment | Control |  | Technology |  | Technology v Control** |  |
| --- | --- | --- | --- | --- | --- | --- |
|  | % | n/N* | % | n/N* | RR (95% CI) | p |
| Any episode of MDP | 47.8% | 928/1941 | 50.7% | 782/1543 | 1.26 (1.12-1.42) | <0.001 |
| Any episode of MDP (first line treatments only)*** | 42.2% | 819/1941 | 48.2% | 744/1543 | 1.30 (1.12-1.52) | 0.001 |
| **Appropriate treatment by first care seeking location (any episode of MDP)** |  |  |  |  |  |  |
| CHW | 49.5% | 146/ 295 | 61.9% | 148/239 | 1.04 (0.86-1.25) | 0.675 |
| Public Facility | 58.0% | 736/1269 | 62.2% | 594/955 | 1.26 (1.07-1.48) | 0.004 |
| Private sector (private facility, pharmacy, shop, herbalist) | 29.4% | 10/34 | 21.7% | 5/23 | 1.66 (0.52-5.34) | 0.392 |
| No care seeking outside of the home | 10.5% | 36/343 | 10.7% | 35/326 | 1.23 (0.75-2.00) | 0.408 |

\*N=total episodes of the described disease; n=total episodes where the correct drugs were received
\*\*adjusted for baseline levels of appropriate treatment for the condition(s) in question, and for variables used to balance arms at baseline (rates of facility and CHW care seeking, cost of care seeking, and CHW motivation score)
\*\*\* First line treatments: artemisinin combination therapy for malaria, ORS for diarrhoea, and amoxicillin for pneumonia

**Table 6b.** Primary Outcome at baseline: Percentage of episodes of MDP appropriately treated

| Outcome | Control | Technology |
| --- | --- | --- |
|  | % (n/N*) | % (n/N*) |
| **Total children in baseline sample** | **2257** | **2165** |
| **% children with MDP\*\*** | 29.3% (662/2257) | 30.4% (658/2165) |
| **Appropriate treatment** |  |  |
| **Any episode of MDP** | **33.1% (298/901)** | **28.2% (261/926)** |
| Suspected malaria | 31.3% (176/562) | 28.4% (158/556) |
| Confirmed malaria | 90.1% (118/131) | 88.7% (110/124) |
| Diarrhoea (ORS) | 56.0% (65/116) | 44.7% (51/114) |
| Diarrhoea (ORS and zinc) | 2.6% (3/116) | 2.6% (3/114) |
| Pneumonia (any antibiotic) | 25.6% (57/223) | 20.3% (52/256) |
| Pneumonia (amoxicillin) | 10.3% (23/223) | 10.6% (27/256) |
| Any episode of MDP at the CHW (first care seeking location) | 35.7% (71/199) | 32.0% (70/219) |
| Any episode of MDP at the Public Facility (first care seeking location) | 46.8% (214/457) | 38.1% (171/449) |
| Any episode of MDP at the Private sector (first care seeking location, includes private facility, pharmacy, shop, herbalist) | 18.8% (3/16) | 19.2% (5/26) |
| Any episode of MDP - no care seeking outside of the home | 10.1% (22/209) | 6.2% (14/226) |

\*N=total episodes of the described disease; n=total episodes where the correct drugs were received. \*\*MDP: denominator includes all children with suspected or confirmed malaria, diarrhoea or pneumonia

**Table 7.** Impact of the inSCALE intervention on appropriate treatment by condition

| Outcome | Control arm | Technology arm |  |  |
| --- | --- | --- | --- | --- |
|  | Freq (%) | Freq (%) | risk ratio versus control (95% CI) | p |
| Appropriate treatment by condition |  |  |  |  |
| Suspected malaria | 54.4% (707/1300) | 56.6% (633/1118) | 1.08 (0.97-1.20) | 0.150 |
| Confirmed malaria | 96.4% (637/661) | 96.2% (589/612) | 1.02 (0.99-1.04) | 0.221 |
| Diarrhoea (ORS) | 29.3% (58/198) | 43.8% (53/121) | 1.76 (1.20-2.56) | 0.003 |
| Diarrhoea (ORS + zinc) | 0.0% (0/198) | 1.7% (2/121) | - | - |
| Suspected pneumonia (any antibiotic) | 36.8% (163/443) | 31.6% (96/304) | 0.80 (0.60-1.08) | 0.142 |
| Suspected pneumonia (amoxycillin) | 12.2% (54/443) | 19.1% (58/304) | 1.50 (1.03-2.17) | 0.033 |

### CHW motivation, identification, clinical knowledge, attrition, and stockouts

The mean motivation and the mean social identification scores were relatively high and did not differ between arms (difference in motivation factors score: -0.03 95% CI -0.57, 0.51; difference in identification factor score: 0.01 95% CI -0.42, 0.43). CHW attrition rates were approximately 18%-19% in both intervention arms with no detected difference (Table 8). Overall scores for CHW clinical knowledge of the signs and management of MDP, also did not differ significantly between arms (Table 8). A breakdown of CHW knowledge scores by condition suggests a trend of improved knowledge in technology arm CHWs particularly in diagnosing pneumonia and diarrhoea - though none reached the threshold for statistical significance and may be chance findings (Table S3 in supplementary file 1). There were no between-arm differences in the rate of stockouts for key MDP treatments in the previous 3 months, though overall rates of stockouts were fairly high at >50% (Table 8).

**Table 8.** Community health worker (CHW) outcomes: Stockouts, attrition, motivation and knowledge in control and intervention arm clusters

| Outcome | Control arm<br>(N = 103) | Technology arm<br>(N = 104) | Effect size (95% CI) | p |
| --- | --- | --- | --- | --- |
|  | Mean (sd) | Mean (sd) |  |  |
| % CHWs experienced a stockout of any MDP drug in last 3 months* | 52.4% (54) | 56.7% (59) | RR: 1.17 (0.92-1.49) | 0.207 |
| Mean CHW motivation score | 4.6 (0.5) | 4.7 (0.5) | RD: -0.03 (-0.57, 0.51) | 0.914 |
| Mean CHW social identification score | 4.7 (0.6) | 4.8 (0.5) | RD: 0.01 (-0.42, 0.43) | 0.976 |
| Mean CHW knowledge score | 19.7 (4.3) | 19.8 (4.4) | RD: 0.97 (-2.01, 3.94) | 0.524 |
| Proportion CHW attrition (%) | 18.4% (13.6) | 18.5% (10.0) | RD: 1.31 (-18.66, 21.30) | 0.878 |

\* Stockouts of any of artemisinin combination therapy, amoxycillin, or ORS. RR- risk ratio; RD- risk difference

### Sensitivity analyses and robustness tests

Sensitivity tests were conducted with two alternative definitions of appropriate treatment: i) only including children with a positive malaria diagnostic test, and ii) including quinine as an appropriate treatment for malaria to account for its use as a second line treatment or during in-patient care (S2 Table in the in Supplementary File 1). Both showed a trend of improved coverage of overall appropriate treatment (20% an 14% improvement, p values 0.006 and 0.073 respectively for i) and ii)). We performed robustness tests of the key primary and secondary impact outcomes appropriate treatment, care seeking, CHW motivation, social identity, clinical knowledge, stockouts, and attrition by repeating the analyses using a comparison of cluster-level mean outcomes (S4 Table in Supplementary File 1). This approach estimated similar significant improvements in appropriate treatment as the random effects model with the exception of the increase in care seeking to CHWs which did not reach significance.

### Meta-analysis of appropriate treatment for MDP: Uganda and Mozambique

Figure 2 shows the results of a fixed effects meta-analysis of the risk ratios (RRs) for percentage coverage of appropriate treatment in Uganda and Mozambique. The analysis suggests implementation of the inSCALE technology model could improve appropriate treatment by 15% (8% - 24%) overall.

**Figure 2.**
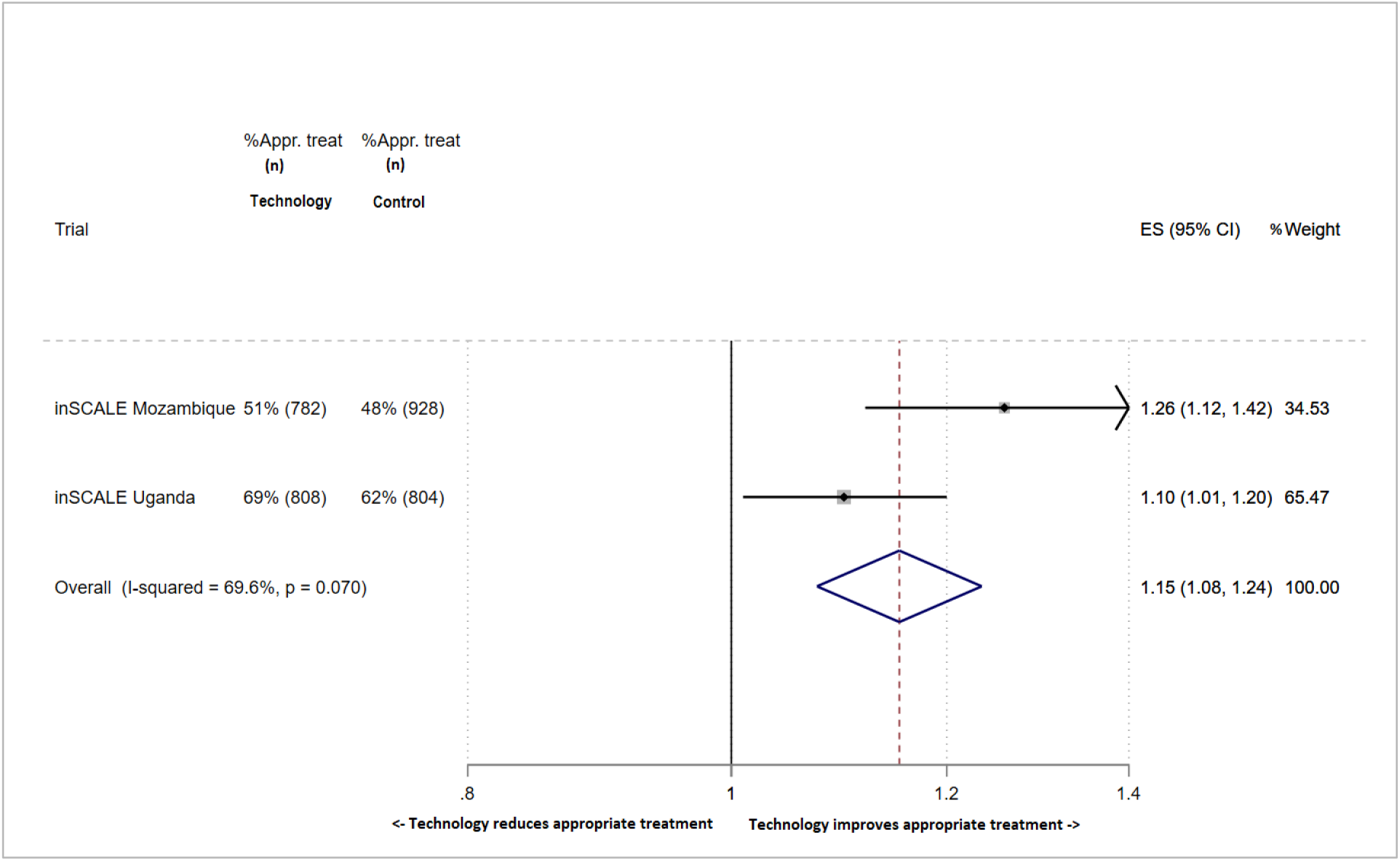
Forest plot displaying an inverse-variance weighted fixed-effect meta-analysis of the effect of the inSCALE technology interventions on appropriate treatment of episodes of suspected malaria, diarrhoea or suspected pneumonia (Uganda and Mozambique). Effect size (ES) is the risk ratio for appropriate treatment. Fixed effects meta analysis (country = fixed effect) Appr. Treat = appropriate treatment of illness episodes (suspected malaria, diarrhoea or suspecter pneumonia). I = intervention arm, C = control arm. ES = baseline-adjusted risk ratio (effect size) of % episodes MDP appropriately treated in intervention versus control arm.

## Discussion

We have demonstrated that a scalable, innovative technology-based intervention implemented in Mozambique can meaningfully improve the coverage of appropriate treatment for common childhood illnesses covered by the iCCM programme by 26-30%. We further showed in a meta-analysis of two countries (Mozambique and Uganda) implementing similar technology-based interventions that an 8-24% improvement in appropriate treatment is likely achievable elsewhere. Our results suggest that iCCM programmes can contribute to the quality of care provided by primary health services by the use of relatively inexpensive technology that prioritises support, efficiency and performance. As a result of this work, the government of Mozambique has expanded the inSCALE technology intervention to seven out of eleven provinces with over 2,500 CHWs and 299 supervisors using the app to date. The programme will be rolled out to all 8,800 CHWs and 1,300 supervisors nationally by end of 2022 and a full data integration plan with DHIS2 is ongoing to optimize strategic decision making(16,17). The expanded programme name, upSCALE, reflects the successful and rapid uptake of our research into national policy and the creation of a national CHW mHealth system(17).

Our conceptual framework for the design of the inSCALE innovation expected that differences in CHW motivation, attrition and social identification would be one of the drivers of improvement in appropriate treatment(15,18). As noted in our formative research and in other settings, ownership of local programmes can be fragmented, CHW interventions are often only tenuously linked with the wider health system and therefore poorly supervised, and their contribution under-recognised(19). The CHWs we surveyed however were in general highly motivated and reported a strong degree of connectedness to the wider health system in both arms at baseline and at the end of follow-up. Local facility records estimated CHW attrition was moderate at 18% over the implementation period and did not differ by intervention arm. Our findings suggest that in contrast to our conceptual framework, alternative routes to impact may be more important in this setting. Though coverage of appropriate treatment in children taken to the CHW was similar across arms, we observed the largest overall increase in coverage of appropriate treatment occurred at public health facilities and in children with diarrhoea, very similar to the pattern of impact observed in the sister inSCALE trial in Uganda [ref]. That these patterns were largely consistent across two countries with separate intervention design and implementation teams is a persuasive indicator of the alternative impact pathways and mechanisms of impact through which inSCALE operates - and warrants further exploration in future studies. This nonetheless likely highlights the integrated nature of inSCALE intervention whereby training and usage of the app by both CHWs and supervising staff at primary care facilities will have increased health worker exposure to guideline-led recommendations for disease management, alerts for low CHW stock for key commodities, motivational messaging, and reporting on prevalence of key childhood conditions. Diarrhoea in particular is often perceived simply as a symptom of a more highly prioritised condition on which treatment is focused(20) and may therefore be overlooked by health workers even if ORS is available (20–22). These findings point to the gains that may be made by adopting an approach that not only strengthens community based iCCM programmes, but the linked primary health care system.

Coverage of the inSCALE innovation after 18 months of follow-up was moderate to good (68% CHWs had a working phone and inSCALE app and 45% had sent a report in the last month). It is difficult to assess this intervention coverage rate in context of similar interventions due to the variability in intervention components, and in endpoint definitions. A SMS (short message service) message-based randomised controlled trial in Tanzania delivered 7 messages to drug shop vendors with recommendations for correct treatment of malaria and reported 70% of dispensers received at least 75% of the intended SMS messages. Whilst this appears comparable to our reported rates, the difference in the health provider type and in the content of the intervention precludes a direct comparison (23). Nonetheless a review of a selection of trials of child health-based mHealth interventions in low and middle income country settings (LMICs) report widely varying intervention coverage rates between 24% and 85% (24,25). Lack of medicinal stock for common childhood conditions has been a barrier to prompt and appropriate treatment in other settings(26), and affected our study sites in Mozambique during and after the period of implementation (8,19). Key issues were the national stock outs of key malaria and antibiotic medications and diagnostic tests within the last 6 months of the study, which may have supressed the full potential impact of the technology intervention for these outcomes. Research by Save the Children in this setting suggests that households who were aware of stockouts at the iCCM CHW may go elsewhere for treatment(19). More than 50% of CHWs across both arms reporting at least one stockout of supplies for the management of MDP in the previous 3 months; such commodity supply barriers were outside the control of our intervention nonetheless it was encouraging that our results highlight a positive impact of the inSCALE app in maintaining or improving overall coverage of clinical management for pneumonia and diarrhoea despite these persistent issues and moderate intervention coverage.

### Wider implications on case prevalence, fatality, productivity, user costs and transmission

The highest case fatality rates for pneumonia, malaria and diarrhoea occur in sub-Saharan Africa(27,28) Separate cost-effectiveness studies for the inSCALE trials in Uganda and Mozambique [**ref**] estimate that increasing coverage of appropriate treatment will have a modest impact on child deaths (1-5%). However important proximal outcomes of such interventions include a potential to reduce community transmission of causative pathogens and/or the risk of recurrent MDP infections (29) and lowering cost of care seeking and increasing equitable access to care (22) through the strengthening and raised profile of iCCM-trained CHWs. Reduced community transmission in particular was suggested by the decrease in prevalence of MDP both over time across the study site in Mozambique, and the additional significantly faster fall prevalence in the intervention arm districts. There is a plausible route to this effect through the increase in the timely use of medications, and/or in awareness of preventative measures (30,31). The reduction in disease prevalence, and improvement in appropriate treatment for diarrhoea and pneumonia are particularly to be welcomed considering no single preventative intervention may be easily applied to control these diseases as a result of their multiple pathogen aetiology(32).

Our study had several limitations. Our estimates of prevalence of MDP and appropriate treatment were based on caregiver reports which may be subject to misclassification. Studies of caregiver recall for pneumonia and malaria symptoms have sensitivities and specificities in the range of 31%-91% depending on the setting(33,34). Recommended strategies to improve the accuracy of disease and treatment estimates include setting the length of the recall period to four weeks (versus 2 weeks used in the inSCALE baseline survey), the use of pictures of available treatments to respondents and the development of survey questions which prioritise local terms and are based on well-validated survey instruments(33–35) including the United Nations- and USAID-supported Multiple Indicator Cluster Survey and Demographic Health Surveys(36,37). We incorporated all these strategies in our surveys and used tailored definitions of disease and treatments in line with national guidelines, including the presumptive use of ACT to treat fever, a practice compounded by nationwide stockouts of iCCM drugs and tests at the time. Our sensitivity analyses of the main outcome with alternative definitions of appropriate treatment [see Table supplementary file 1] nonetheless produced similar results – of note, restricting cases of fever to those with a confirmed malaria test estimated a 20% improvement in appropriate treatment which remained highly significant. The inSCALE study evaluated the impact of strategies to improve scale-up and coverage of iCCM programmes using a randomised controlled design as we believed this method would provide the most convincing effectiveness estimates for stakeholders. The number of district clusters available for randomisation was however determined by iCCM programme readiness (iCCM-trained CHWs in place) and lack of concurrent interventions in the area at baseline. This resulted in twelve districts available for randomisation. We tested the robustness of our random-effects models with cluster-averaged t-tests of our primary outcomes as recommended by Hayes and Moulton(38), and whilst less powerful, we observed similar estimates of impact on appropriate treatment (See supplementary file 1).

## Conclusions

The body of research evidence for digital and mHealth programmes is increasing, however there remains a significant gap for evidence from good quality studies of the potential for available and emerging technologies to transform health outcome in LMICs for children, particularly when delivered via community-based health worker cadres(39). Mobile phone penetration is predicted to increase in sub-Saharan Africa(40) and is estimated to be a key medium by which health care access and service delivery can be improved for the underserved. The WHO’s recently published set of global recommendations for digital interventions for health services delivery highlight the potential of mHealth interventions for a range of uses and includes the key components of the inSCALE technology intervention – clinical decision support, stock tracking and provider-to-provider telemedicine (41). Significantly, the 2019 WHO/UNICEF iCCM strategy meeting in Addis Ababa recommended “Interventions to improve quality, including supportive supervision and mentoring of CHWs in designated health facilities, are essential to ensure high-quality iCCM and should be budgeted for and included in district plans”(42). This renewed encouragement for implementation of iCCM this decade may encourage relevant stakeholders looking to efficiently scale up programmes to explore digital solutions for their context. Our study and meta-analysis together with the expansion of the inSCALE programme in Mozambique (16,17) highlight the potential scalability of an mHealth intervention aimed at strengthening iCCM systems to address the largest causes of childhood morbidity and mortality in sub-Saharan Africa.

## Materials and Methods

### Study Site

The study took place in the Inhambane Province of southern Mozambique between November 2013 and July 2016. The province covers a geographical area of 68,775km^2^ with a population of approximately 1.4 million of which 17% are under 5 years of age(43). The Inhambane mortality rate in this age group at the start of the study was 76.4 deaths per 1000 livebirths(44), with more than 60% of all deaths in the province caused by a combination of malaria, diarrhoeal diseases and pneumonia(45,46). Sine 2011, iCCM had been included in the Inhambane programme of work for Agentes Polivalentes Elementares (APE), the name for the local cadre of community health worker and referred to as CHWs in this paper. Government and donor funding for the programme supported the training, supervision and provision of iCCM commodities for all CHWs in the province throughout the study period. A detailed study protocol for the inSCALE trial in Mozambique has been published elsewhere(15). Details of the setting and study design for the inSCALE study in Uganda, used for the metanalysis presented in this paper, is published elsewhere [**ref**].

### Study Design and Participants

We conducted a superiority open label two-arm cluster-randomised controlled trial in 12 districts in Inhambane Province in southern Mozambique. Six districts (Funhalouro, Govuro, Jangamo, Panda, Vilankulo, and Zavala) were randomised to the inSCALE intervention and six (Homoine, Inharrime, Inhassouro, Mabote, Massinga, and Morrumbene) to the control arm. The provincial and economic capitals of Inhambane, Inhambane and Maxixe respectively, was excluded from the randomisation and Maxixe was instead used for piloting of the intervention in its rural areas during the design stage. Intervention allocation was randomised at the district level as this was the lowest unit of administration and training for the CHW iCCM programme and therefore for implementation of the inSCALE intervention. Eligible households within the district were defined as those with at least one resident child under five years of age at any point during the study in a community served by at least one iCCM-trained CHW. Local data collected by Malaria Consortium in the area in 2011 suggested that 45% of households within each community fit the eligibility criteria.

Ethical approval for the study was obtained from the Mozambique National Bioethics Committee for Health (reference 345-CNBS-10) and the London School of Hygiene & Tropical Medicine Ethics Committee in the UK (reference 5762). Approval for the random allocation of districts to intervention or control groups and implementation of the interventions was obtained from the Inhambane Provincial Office responsible for health administration for the province. Written informed consent was obtained from the main caregiver of each child under 5 years of age for baseline and follow up data collection. Consent forms included an explanation of the purpose of the data collection and confirmed participants were free to withdraw at any time. The inSCALE trial is registered on clinicaltrials.gov with identifier NCT01972321.

### Randomisation and Blinding

In order to reduce the likelihood of chance imbalances between intervention arms at baseline given the number of clusters available for randomisation(38,47)., we performed a randomisation that was restricted with respect to the percentages of households with sick children seeking care at a public health facility, the percentage seeking care to a CHW, average CHW motivation (composite) score and the log cost of care seeking for sick children. Data for these parameters was obtained from a baseline survey conducted between November-December 2012. We used the statistical software Stata (version 13) to generate the 924 possible randomisation schemes for the 12 districts and picked one at random from the subsection of 84 schemes that fitted our balance criteria(48) [see online supplement file 1]. Due to the nature of the technology intervention, it was not possible to blind participants or fieldworkers to the intervention allocation, however allocation was masked in the database and all analyses were initially performed on data with intervention allocation removed and repeated on unblinded data after agreement on the final analysis methods with study partners.

### Study Procedures – data collection

The baseline survey was conducted in all households in 2-8 randomly-selected communities (known as enumeration areas or EAs) sampled proportional to size in each district in order to cover approximately 500 households per district (mean of 91 households per village), which would provide 80-90% power to detect an impact of the intervention in accordance with the estimated sample sizes for detecting intervention impact at endline as described in the inSCALE protocol paper(15). Approximately 21 CHWs were also surveyed per district at baseline (1-2 per village). Community sampling frame data were based on complete enumeration information provided by the District Offices and EA location, size and households were verified by field workers.

The baseline survey was administered to caregivers of children under 5 years of age, and collected information on symptoms of malaria, diarrhoea, and pneumonia (MDP) in these children in the last 4 weeks, care seeking for these illnesses, costs of care, socioeconomic data at the household level. Household questionnaires were based on standard health survey instruments used in the country (Mozambique DHS(36) and Malaria Indicator Cluster Surveys(37)). The CHW survey included data on CHWs’ clinical knowledge, time use, motivation and ‘social identity’ – or sense and degree of connectedness to the local and national CHW and health services systems. The CHW clinical knowledge instrument was developed based on the format and example clinical event scenarios in the WHO Health Facility Survey Tool template (49). The likert scale-based CHW motivation and identity tool used a pictorial scale indicating degree of agreement with statements about enthusiasm for the job, motivation and identification with the local and national iCCM programme, and was developed based on extensive formative research by the study team on health worker motivation and social identity (50,51). Both tools were adapted to the Mozambique setting prior to usage with the input of facility staff and CHWs in an iterative process that developed wording, accompanying graphics and reference to local health system structures and clinical guidelines.

Household and CHW surveys were also extensively field piloted with field staff including workshops role playing and refining of translations, and all tools were delivered in Portuguese. A quality control check of 10% of the weekly survey households was undertaken by field worker supervisors. Pictures of locally available medications were used in the household surveys to improve the accuracy of reports of treatment for MDP(33).

At the end of the follow-up period an endline impact survey was conducted in May-June 2015 in the same villages visited at baseline in households with children meeting inclusion criteria at endline, and again covering all active CHWs serving the selected villages. The survey collected data across the same themes to estimate study impact and included additional questions to CHWs and caregivers on coverage of the inSCALE innovation components in the intervention arm. All household and CHW data collection tools are available in Supplementary File 2.

### Intervention

The inSCALE technology intervention was developed in collaboration with Dimagi a social enterprise corporation focused on the development of digital solutions for health workers in low and middle income settings (52). Steering and input was received from the Mozambique ministry of health, community and facility-based stakeholders, funding bodies and the inSCALE trial steering committee(15,53). The aim of the innovation was to leverage technological strategies to strengthen links between fragmented CHW programmes and the primary care health services including links between CHW peers and health facility staff, and by doing so to improve CHW outcomes and child health outcomes for children. The inSCALE technology intervention development process and conceptual framework linking intervention components, outputs and outcomes is described in detail in the protocol paper (15). The technology intervention was implemented in July 2013, and after a 6 month roll-out and embedding period, follow-up to impact analysis continued to July 2015 (18 months). iCCM-trained CHWs and their health facility supervisors in the intervention arm were provided with a Samsung® smartphone installed with the inSCALE mHealth application, a solar charger with a number of pin adapters, and a solar lamp for night-time consultations.

Intervention arm CHWs and health facility supervising staff were trained in the use of the mHealth tools. Training was conducted using a cascade approach where 7 master trainers trained 18 facility- and community-based trainers who in turn trained 47 health facility supervisors and 132 CHWs in the technology arm over 4 days with follow-up support provided by the project team during the implementation period. The inSCALE mHealth mobile app consisted of a sick child decision support algorithm in a user-friendly interface including multimedia, audio and images to improve adherence to iCCM treatment protocols. CHWs were additionally able to use the phones to track consultations with sick children, make diagnoses and management choices, report key statistics to a central server, request stock from their supervising facilities, and maintain a register of patients seen. These reports were forwarded on a weekly basis to health facility supervising staff who received tailored follow-up supervision prompts. The inSCALE app also worked offline, therefore where there were network coverage issues, CHWs and health facility supervising staff had the option to delay sending of reports until they were in a location with better coverage. CHWs and supervisors were allocated to local closed user groups which enabled complimentary calls with the inSCALE smartphones to be made without charge to fellow members in the group. Finally, CHWs and their health facility supervisors received weekly (CHWs only) and monthly (CHWs and facility supervisors) automated motivational messages tailored to their report results with the tone and emphasis based on formative work. After an initial 6-month roll-out and embedding phase, the technology intervention continued to be implemented in intervention arm districts for an additional 18 months. Standard care during this period was available in both intervention and control arm districts, where CHWs and health facilities provided guideline-led management for childhood illnesses via the underlying iCCM programme and integrated management of neonatal and childhood illnesses (IMNCI) services(9,54) respectively.

Data on CHW recruitment, training, resignations, and replacements were tracked by supervising health facilities at district level and this data was regularly extracted into the inSCALE database during the follow-up period by the study team in order to estimate attrition rates.

### Outcomes

The primary outcome for the trial was the percentage of recent cases of MDP in the last 4 weeks in children 2 months to 5 years of age that were treated using appropriate medication(15). Secondary outcomes were the prevalence of MDP in children at endline, the percentage of cases of MDP appropriately treated with recommended first line medications, the percentage of children who sought care for MDP by provider type, the percentage of CHWs who had had a stock-out of iCCM medications in the past 3 months, CHW attrition rate calculated as the proportion of CHWs who left their post during the follow-up period out of the total CHWs trained at baseline, CHW clinical knowledge scores (overall and by condition subgroup), and composite scores describing CHW enthusiasm and drive for the job (motivation) and sense of connectedness to the local and national iCCM programme (social identification). CHW knowledge scores were calculated as the sum of correct clinical decisions mentioned by the CHW in response to a series of scenarios involving children with symptoms of mild and severe disease. Points for correct items were weighted according to an algorithm defined in Supplementary file 1. CHW motivation and social identification scores were calculated by a factor analysis and empirical item reduction process applied to original field tool responses. The methods have been detailed previously(55), and resulted in a three-factor CHW motivation scale of 17 items measuring general motivation, feelings of reward for effort, and retention in role. An additional single factor CHW scale of 4 items was identified which explained the majority of the variation in social identification between CHWs. Retained items from both the motivation and social identification tools showed good internal consistency (alpha = 0.67 for CHW motivation and alpha = 0.69 for CHW social identification). The final motivation scores were calculated as the product of the rotated factor analysis regression coefficients and the Likert value for each original item. The final social identification scores were calculated as the sum of the original 4 items (after reverse coding as appropriate).

Additional impact measures included the percentage of children who were appropriately treated overall whether they had symptoms of one or more of MDP in the last 4 weeks (i.e. a per-child analysis), and two sensitivity analyses of the main impact outcome using alternative definitions of appropriate treatment. All definitions of appropriate treatment and medications used in primary secondary and additional endpoints are provided in Supplementary File 1 (Table S8).

### Statistical analysis

All analyses were based on intention-to-treat principles and accounted for the cluster-randomised design. Multivariate mixed-effects models were specified for all impact outcomes, which account for the clustering of outcomes by inclusion of a random intercept at the district level. Intervention arm, the cluster mean percentage of children who were appropriately treated at baseline, and the parameters used to balance intervention arms during randomisation were included as fixed effects. Unadjusted analyses of impact were also performed, and the results included in supplementary file 1 (Table S1). For the primary outcome and other binary models, odds ratios were converted to risk ratios using the marginal standardisation technique and 95% CIs estimated using the delta method (56). Additional sensitivity analyses of the primary outcome were conducted to account for alternative treatments available for MDP [see Supplementary file 1 Table S8 for definitions]. Robustness checks of the primary outcomes were conducted by estimating cluster averaged results using t-tests following the procedures of Hayes and Moulton(38), recommended when the number of clusters per arm is less than 10(38) [see Supplementary file 1]. CHW attrition was estimated using cluster-averaged mean rates as human resources information was provided at this level only from district health offices. In order to estimate a pooled effect of the inSCALE technology intervention on appropriate treatment across both Uganda and Mozambique, we performed a meta-analysis of effect sizes by combining the results of the Mozambique (this study) and Uganda [**ref**] trials of the inSCALE intervention. As there was no significant heterogeneity (<50% estimated from exploratory meta-analysis models) between studies, we present the pooled result from a fixed effects meta-analysis.

## Data Availability

Data will be available after publication at this address:
https://doi.org/10.17037/DATA.00002559

## Supplementary file Tables

| Table | Caption |
| --- | --- |
| **Supplementary file 1** |  |
| S1 | Unadjusted analyses of primary outcomes |
| S2 | Sensitivity analyses for primary outcome at endline (any episode of MDP): Alternative definitions of appropriate malaria treatment |
| S3 | Whole Child Analysis – Appropriate treatment |
| S4a | Robustness tests (cluster-averaged outcomes) – Primary Outcomes |
| S4b | Robustness tests (cluster-averaged outcomes) – Secondary Outcomes: appropriate treatment stratified by condition or provider |
| S5 | Robustness tests (cluster-averaged outcomes) – Secondary outcomes: prevalence of suspected and confirmed malaria, malaria blood testing, diarrhoea, and pneumonia |
| S6 | CHW clinical knowledge by subgroup (diagnosis of MDP, treatment of MDP) |
| S7a | Item weighting – Case study: Francis/Alberte (pneumonia and diarrhoea) |
| S7b | Item weighting – Case study: Hope/Tina (malaria) |
| S7c | Item weighting – Case study: Beatrice/Janete (severe pneumonia) |
| S7d | Item weighting – Case study: Muteesa/Kizito (severe disease/malaria) |
| S7e | Item weighting – Case study: James (severe disease/malaria) |
| S7f | CHW Knowledge scoring for subcategories: diagnosis and treatment of suspected malaria, diarrhoea and pneumonia |
| S8 | Definitions of appropriate treatment and other key child health outcomes |
| **Supplementary file 2** |  |
| inSCALE data collection forms |  |

## Contributors

SS, KK, RL, FK, ACB, AB, DS, ZH, GA, JT, NS, JC, FB, SM and BK designed and performed the study. SS analysed the data. SS, RL and KK drafted the manuscript and FK, ACB, AB, ZH, GA, JT, NS, JC, FB, SM and BK reviewed the manuscript. KK, RL, FK, DS, ZH, JT provided critical comments on the review of the manuscript. All authors have read and approved the final manuscript.

## Availability of study data

The data for the inSCALE project is available on the public repository LSHTM Data Compass under DOI https://doi.org/10.17037/DATA.00002559.

## Acknowledgements

We wish to thank the local research team who collected and managed the data, diMaggi for support in designing the app software, the CHWs and supervisors who delivered the intervention, and the participants of the study for their time and insight. Funding for this research was provided by the Bill & Melinda Gates Foundation [grant code OPP1002407] and UK Department for International Development (UK Aid) through grants to Malaria Consortium, the London School of Hygiene and Tropical Medicine, and the Institute for Global Health, University College London for the inSCALE project (http://www.malariaconsortium.org/inscale/). The funders participated in the initial discussions on intervention design but had no role in the design of the evaluation, data gathering, analysis, interpretation, or writing up of the findings. SS, KK, RL, NB, FK and BK had full access to all the data and SS had final responsibility for the decision to submit for publication.

